# Aligning SARS-CoV-2 Indicators via an Epidemic Model: Application to Hospital Admissions and RNA Detection in Sewage Sludge

**DOI:** 10.1101/2020.06.27.20141739

**Authors:** Edward H. Kaplan, Dennis Wang, Mike Wang, Amyn A. Malik, Alessandro Zulli, Jordan Peccia

## Abstract

Ascertaining the state of coronavirus outbreaks is crucial for public health decision-making. Absent repeated representative viral test samples in the population, public health officials and researchers alike have relied on lagging indicators of infection to make inferences about the direction of the outbreak and attendant policy decisions. Recently researchers have shown that SARS-CoV-2 RNA can be detected in municipal sewage sludge with measured RNA concentrations rising and falling suggestively in the shape of an epidemic curve while providing an earlier signal of infection than hospital admissions data. The present paper presents a SARS-CoV-2 epidemic model to serve as a basis for estimating the incidence of infection, and shows mathematically how modeled transmission dynamics translate into infection indicators by incorporating probability distributions for indicator-specific time lags from infection. Hospital admissions and SARS-CoV-2 RNA in municipal sewage sludge are simultaneously modeled via maximum likelihood scaling to the underlying transmission model. The results demonstrate that both data series plausibly follow from the transmission model specified and provide a 95% confidence interval estimate of the reproductive number *R*_0_ ≈ 2.4 ±0.2. Sensitivity analysis accounting for alternative lag distributions from infection until hospitalization and sludge RNA concentration respectively suggests that the detection of viral RNA in sewage sludge leads hospital admissions by 3 to 5 days on average. The analysis suggests that stay-at-home restrictions plausibly removed 89% of the population from the risk of infection with the remaining 11% exposed to an unmitigated outbreak that infected 9.3% of the total population.

Highlights
- A maximum likelihood method for aligning observed lagged epidemic indicators via an underlying transmission model is derived and illustrated using observed COVID-19 hospital admissions and SARS-CoV-2 RNA concentrations measured in sewage sludge to model a local SARS-CoV-2 outbreak
- The method enables direct estimation of the reproductive number *R*_0_ from the observed indicators along with the initial prevalence of SARS-CoV-2 infection in the population at risk
- The analysis suggests tracking SARS-CoV-2 RNA concentration in sewage sludge provides a 3 to 5 day lead time over tracking hospital admissions, consistent with purely statistical time series analysis previously reported
- The model enables estimation of the fraction of the population compliant with government-mandated stay-at-home restrictions, the size of the exposed population, and the fraction of the population infected with SARS-CoV-2 over the outbreak

## 1 Introduction

Ascertaining the state of coronavirus outbreaks is crucial for public health decision-making. Absent repeated representative viral test samples in the population (Kaplan and Forman 2020), public health officials and researchers alike have relied on lagging indicators of infection to make inferences about the direction of the outbreak and attendant policy decisions. How useful these indicators are depends upon their typical lags behind the incidence of infection. Some indicator lags, such as time from infection to hospitalization, have been studied empirically (Lewnard et al 2020, CDC 2020, MIDAS 2020). Other indicators have been proposed with the hope that they would greatly reduce the lag time from infection. One such promising indicator is measured SARS-CoV-2 RNA concentration in municipal wastewater (Foladori et al 2020, Hart and Halden 2020, Peccia et al 2020). How much earlier might such a signal inform officials of changes in the state of the outbreak?

This paper tackles this question by using an epidemic transmission model to create model-scale versions of whatever indicator is of interest, and then scales these model quantities to match observed indicator values in the real world. This approach clarifies the time lags that should be expected from SARS-CoV-2 incidence to whichever indicator is of interest, and by doing so makes it possible to compare the relative timing of one indicator to another, providing the model fit to the data is sufficiently close.

Our study takes advantage of recently conducted research tracking the local SARS-CoV-2 outbreak in the New Haven, Connecticut, USA metropolitan area. As reported by Peccia et al (2020), daily SARS-CoV-2 RNA concentrations were obtained by sampling sewage sludge from the local wastewater treatment plant and conducting PCR tests to determine virus RNA concentration. Daily COVID-19 admissions to the Yale New Haven Hospital restricted to residents of the same four towns served by this wastewater treatment plant were also recorded over the same time period. An epidemic model developed by Kaplan (2020b) was taken as the basis for calibrating these two lagging indicators while simultaneously estimating the initial condition and reproductive number *R*_0_ of this outbreak. This paper details the methodology employed and results obtained from doing so.

The next section presents a quick description of the transmission model reported in Kaplan (2020b). In Section 3, a simple method is described for linking model-scale lagging epidemic indicators to SARS-CoV-2 incidence based on the model and appropriately defined lag probability density functions, which enables a model-scale comparison of different indicators to see how they should appear over the course of an outbreak (Figure 1). Section 4 presents a simple statistical approach to analyzing real-world indicator data by scaling modeled indicators up to observed values based on maximum likelihood estimation while also estimating the initial condition and reproductive number of the epidemic wave from the underlying transmission model. We simultaneously scale hospital admissions and the RNA virus concentration observed in the sewage sludge to the epidemic model (Table 1, Figure 2 and 3). The results show that accounting for the inherent noise in the data, both the virus RNA concentration in the sewage sludge and hospital admissions match the model expectations reasonably well, and provides a 95% confidence interval for the reproductive number *R*_0_ ≈ 2.4 ± 0.2. Section 5 reports a sensitivity analysis to allow for different probability distributions for the lags from infection to hospital admissions and sludge RNA concentration respectively. The analysis verifies that there is a 3 to 5 day separation between the sludge RNA concentration and hospital admissions curve, consistent with earlier analysis based on statistical time-series analysis (Peccia et al 2020). Section 6 uses the preceding analysis to provide epidemic insights suggesting that stay-at-home restrictions effectively bifurcated the local population by plausibly removing 89% of the population from the risk of infection with the remaining 11% exposed to an unmitigated outbreak that infected 9.3% of the total population. Section 7 provides a summary of the key points of the paper.

**Figure 1:**
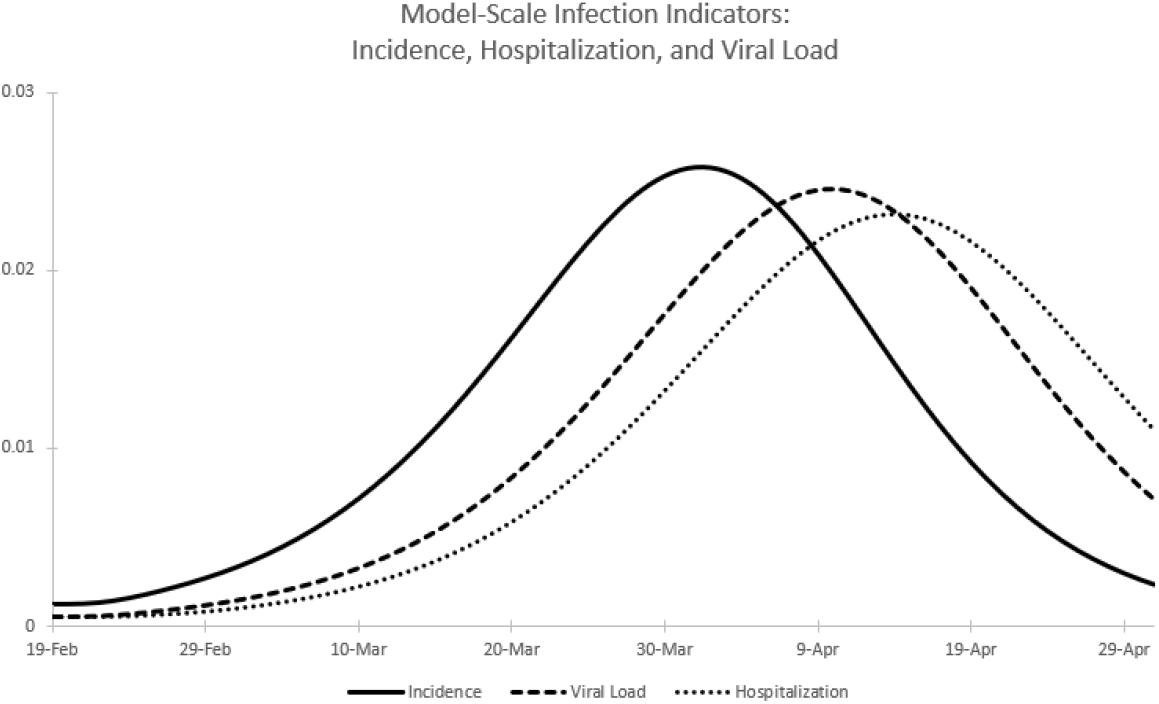
Model-scale infection indicators (all units in infections per person per unit time): SARS-CoV-2 incidence (solid line), sludge viral load (dashed line), hospital admissions (dotted line).

**Table 1.**

| Parameter | Maximum Likelihood Estimate | Standard Error |
| --- | --- | --- |
| $\pi(0)$ | 0.0161 | 0.0032 |
| $k_H$ | 1006.603 | 56.847 |
| $k_V$ | 57.589 | 4.867 |
| $c_V$ | 12.890 | 2.951 |
| $R_0$ | 2.383 | 0.100 |

**Figure 2:**
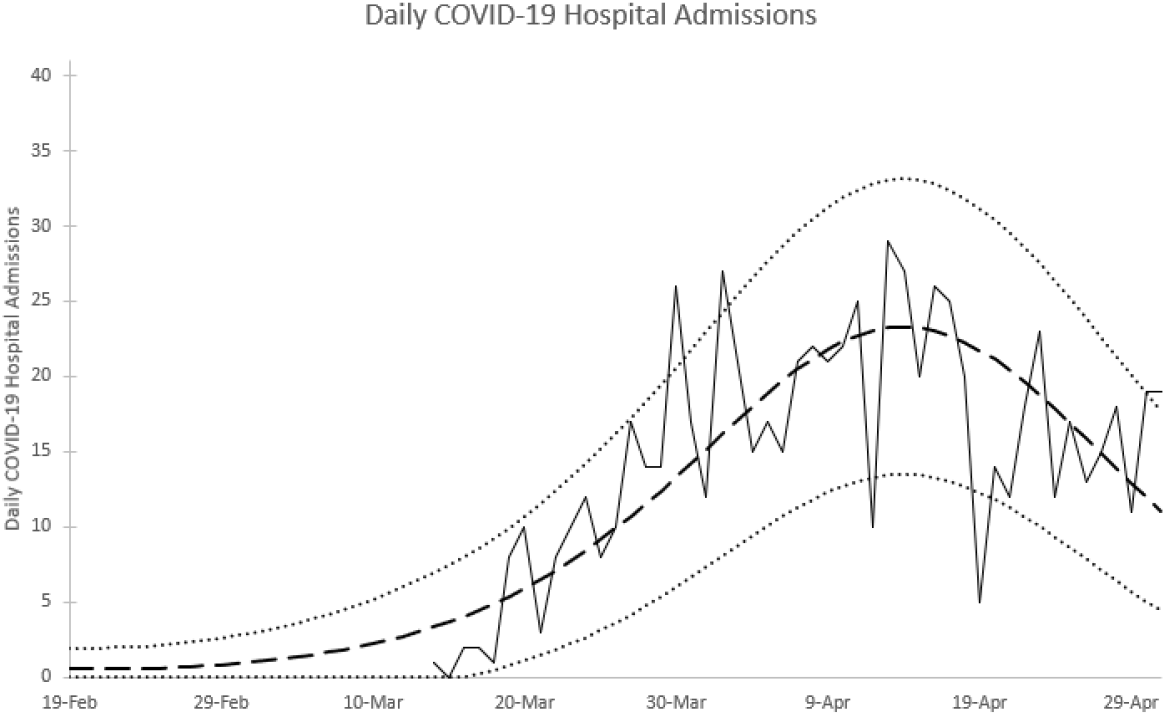
Daily COVID-19 hospital admissions: observed data {solid line), model-based expected value (dashed line), 95% prediction interval limits (dotted line).

**Figure 3:**
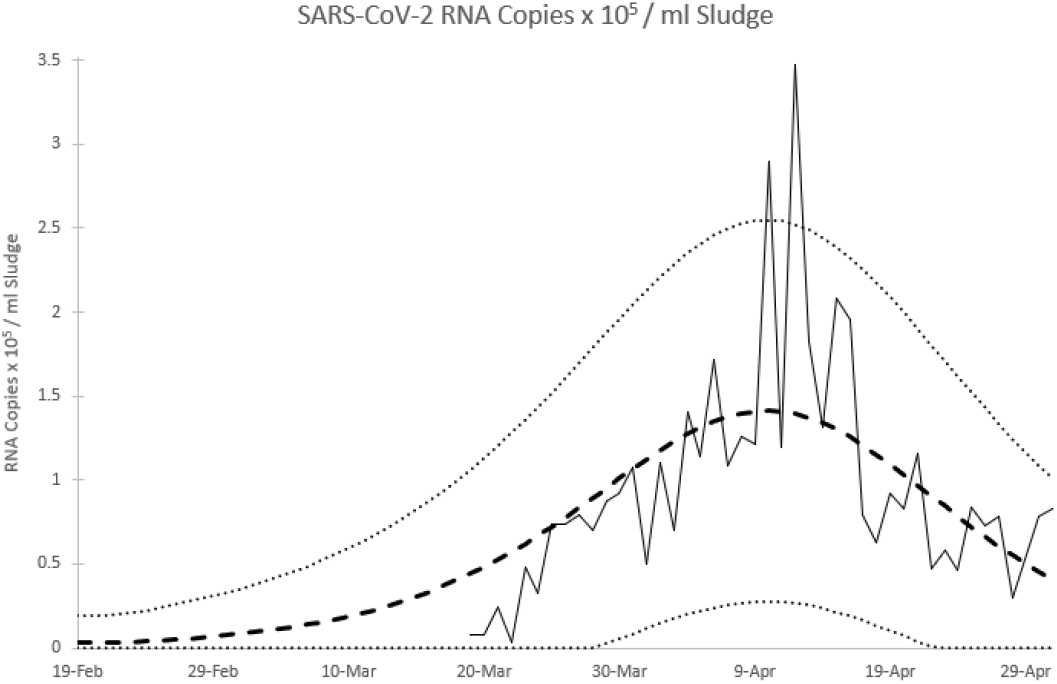
SARS-CoV-2 RNA Copies x 105 / ml Sludge: observed data (solid line), model-based expected value (dashed line), 95% prediction interval limits (dotted line).

## 2 Transmission Model

Data detailing person-to-person SARS-CoV-2 transmission in Wuhan were reported by Li et al (2020). These data enabled an early model-based assessment of prospects for containing coronavirus via isolation and quarantine (Kaplan 2020a), while that analysis was extended to a dynamic transmission model for SARS-CoV-2 transmission in Connecticut (Kaplan 2020b). This latter model incorporates infection-age-dependent transmission, and thus falls into the class of renewal equation epidemic models (Heesterbeek and Dietz 1996, Champredon and Dushoff 2015). The key model element is the age-of-infection dependent transmission rate λ(*a*), which can be thought of as the instantaneous transmission intensity of an individual who has been infected for *a* time units. At the beginning of an outbreak when an infectious person is embedded in an otherwise susceptible population, the expected number of infections transmitted per infectious person equals the reproductive number *R*_0_, which is given by

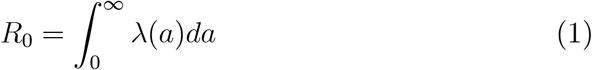

as is well known. Li et al (2020) reported estimates of both the exponential growth rate *r* and backwards generation time probability density function *b*(*a*), enabling λ(*a*) to be written as

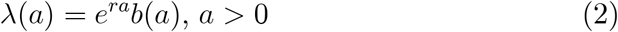

(Kaplan 2020a, 2020b; Britton and Tomba 2019; Champredon and Dushoff 2015; Wallinga and Lipsitch 2007), which together imply a point estimate of *R*_0_ =2.26(Kaplan 2020a), consistent with values widely reported elsewhere (Ferguson et al 2020, Kissler et al 2020, MIDAS 2020, Park et al 2020).

An alternative representation of λ(*a*) is

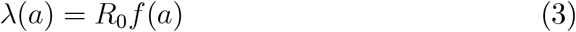

where *f*(*a*) is the *forward* generation time density that dictates the timing of transmission (Britton and Tomba 2019; Champredon and Dushoff 2015; Wallinga and Lipsitch 2007). We adopt this representation in the present analysis, as it enables estimation of the underlying reproductive number *R*_0_ directly from the data at our disposal.

The transmission model developed in Kaplan (2020b) that will be used to anchor our infection indicators analysis follows. Let

*ψ*(*t*) ≡ transmission potential (or force of infection) at chronological time *t*;

*s*(*t*) ≡ fraction of the population that is susceptible to infection at chronological time *t*;

*π*(*a,t*) ≡ density of the population that has been infected for duration *a* at time *t*;

*π*(0,*t*)= incidence of infection at time *t*.

Given the initial condition *π*(*a*, 0) which implies 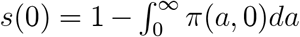, the model equations are:

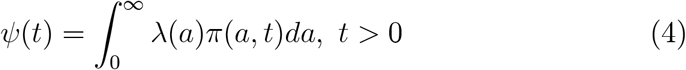

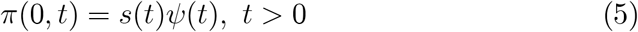

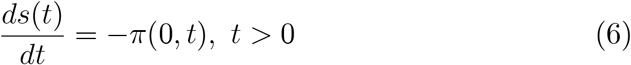

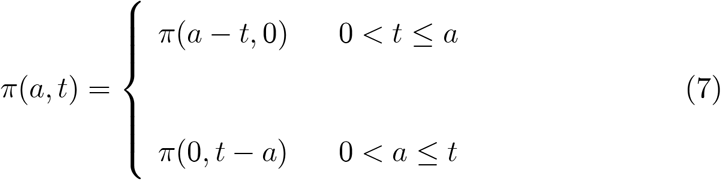

Equation (4) defines the transmission potential at time *t*, which is infection-age-dependent transmission λ(*a*) weighted by the infection-age-dependent prevalence of infection in the population *π*(*a,t*);equation (5) equates SARSCoV-2 incidence to the product of the fraction of the population that is susceptible and the transmission potential; equation (6) depletes susceptibles with the incidence of infection; and equation (7) aligns the fraction of the population infected for duration *a* at time *t* with the incidence of infection at time *t* − *a*, adjusting for the initial conditions at time zero. There is no additional accounting for the duration of infectiousness because the time course of infection is already built into λ(*a*).

The *final size ϕ*, defined as the fraction of the population that is infected over the duration of an outbreak described by this model, follows (Kaplan 2020b)

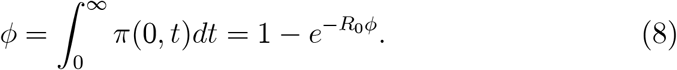

We will make use of this relationship below.

The transmission function employed in our base case analysis is given by equation (3) using the forward generation time density *f*(*a*) implied by Li et al (2020), which is a gamma density with mean (standard deviation) equal to 8.86 (4.02) days (see Figure 4b). The reproductive number *R*_0_ and initial conditions *π*(*a*, 0) and hence *s*(0) are estimated from the data as described below.

**Figure 4:**
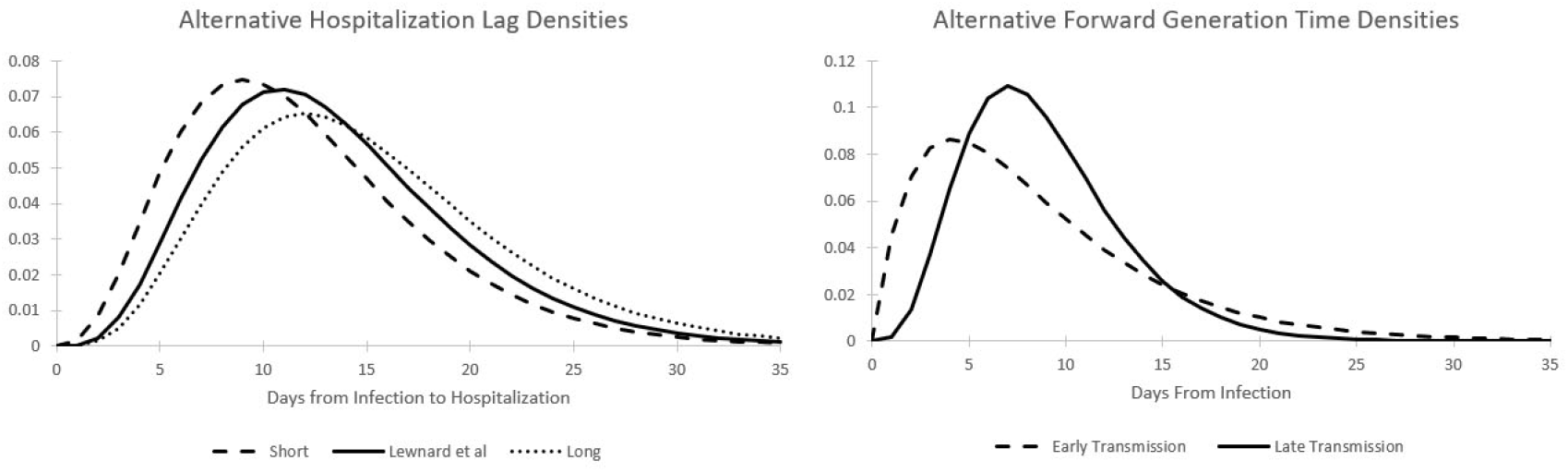
a. Hospitalization lag density functions indicating short (mean 12 days), Lewnard et al (mean 13.5 days), and long (mean 15 days) lags. b. Forward generation time densities indicating early transmission (Park et al) and late transmission (Li et al) of infection.

## 3 Model-Scale Infection Indicators and Time Lags

In the absence of repeated representative viral testing in a population, officials and researchers alike have turned to lagging indicators of infection such as diagnosed COVID-19 cases, hospitalizations, and deaths to monitor the state of the outbreak. How useful such indicators are depends upon their lag time from infection. Let *y*(*t*) be the value of a model-scale infection indicator that represents a distributionally lagged signal of the incidence of infection. Specifically, denote *L_Y_* as the time lag from infection, and define 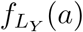 as the probability density function governing the lag *L_Y_*. The model-scale infection indicator *y*(*t*) is then defined as

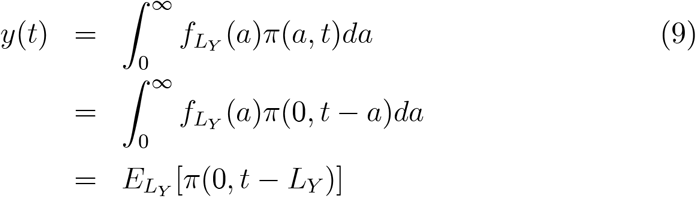

where *E_X_* [·] denotes mathematical expectation with respect to random variable *X*. A first-order Taylor approximation yields the approximation

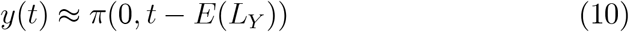

which suggests that the model-scale indicator value can be approximated by incidence evaluated *E*(*L_Y_*) time units earlier. The model-scale indicator at time *t* is just the expected value of SARS-CoV-2 incidence *L_Y_* time units into the past. Note from equation (8) that

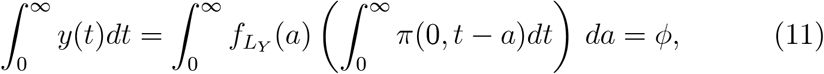

which shows that the model-scale indicator solely reflects the *timing* at which SARS-CoV-2 incidence is experienced for whatever indicator is of interest while conserving the total incidence of infection. The units for all modelscale indicators thus equal infections per person per unit time, regardless of which real-world indicator is being considered.

### 3.1 Example: Hospital Admissions

Hospital admissions have been used as an indicator for the coronavirus outbreak under the presumption that the fraction of new infections that require hospitalization remains constant over time. Define *L_H_* as the time from infection to hospitalization for those infected persons that do require hospital treatment. A review of several published studies by Lewnard et al (2020) estimated that the time from infection until hospitalization averages 13.5 days with 95% probability coverage ranging from 4.8 to 27.9 days. We approximate this finding by employing a gamma distribution with *α* = 4.954 and *β* = 2.725 to represent the probability density of *L_H_*, 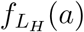. This distribution also has a mean of 13.5 days with 95% probability coverage ranging from 4.4 to 27.7 days (see Figure 4a). Similar times from infection to hospitalization are implied by the Centers for Disease Control COVID-19 pandemic planning scenarios (CDC 2020) and also MIDAS (2020). We employ this density in our base case analysis, but will consider distributions with shorter and longer times from infection to hospital admissions in the sensitivity analyses of Section 5.

Conditional upon the transmission model described in equations (4-7), the model-scale hospitalization indicator *h*(*t*) is, following equation (9), given by

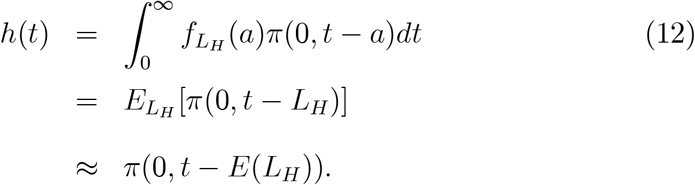

Figure 1 plots both the model-scale SARS-CoV-2 incidence *π*(0,*t*) and hospitalization indicator *h*(*t*) assuming λ(*a*) as defined in equation (3) with *R*_0_ = 2.38 as will be estimated subsequently; the gamma distribution for 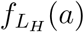 described above; *s*(0) = 1 − 0.0161 reflecting the initial prevalence of infection as estimated below (with time 0 taken as February 19, 2020); and *π*(*a*, 0) = 0.0161/30 for 0 < *a* ≤ 30 as explained below. The model-scale hospitalization indicator lags incidence by about two weeks, as one would expect given that *E*(*L_H_*) = 13.5 days by design.

### 3.2 Example: SARS-CoV-2 RNA in Municipal Sewage Sludge

Peccia et al (2020) reported daily SARS-CoV-2 RNA concentrations based on sampling sludge from a municipal wastewater treatment plant serving the combined 200,000 population of the towns of New Haven, East Haven, Hamden, and Woodbridge in the state of Connecticut, USA. Virus RNA concentrations in sludge should reflect the amount of virus shed in feces by infected persons in the population served by the treatment plant, resulting in a fecal estimate of community virus RNA concentration. Though virus RNA concentrations in feces degrade exponentially with the time from excretion to sample collection (Foladori et al 2020, Hart and Halden, 2020), virus RNA concentrations obtained from sludge sampled daily should be discounted by approximately the same degradation factor, rendering the resulting signal a plausible surrogate tracking community virus RNA concentration over time.

Referring back to the epidemic model, the appropriate measure of virus RNA concentration is the transmission potential *ψ*(*t*), as the amount of virus shed in feces should reflect the average infectiousness of the population. However, to use the indicator framework developed above, the age-of-infection transmission rate λ(*a*) must be normalized to the scale of a probability density function. This is easily achieved by defining

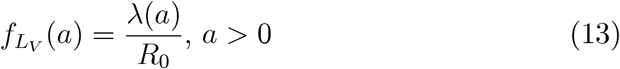

which is immediately recognized as the forward generation time probability density *f*(*a*) introduced earlier. This density enables the definition of the model-scale virus RNA indicator *υ*(*t*) as

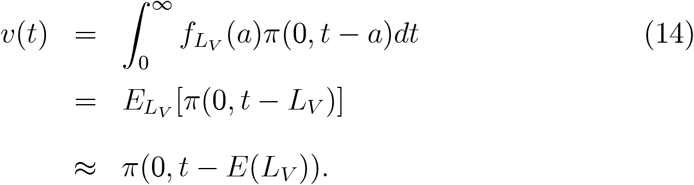

For λ(*a*) as defined in equation (3) using the forward generation time density *f*(*a*) corresponding to Li et al (2020), the expected lag *E*(*L_V_*) is given by 8.9 days, or 4.6 days shorter than the lag from infection to hospitalization (we consider an alternative generation time distribution in Section 5). Figure 1 reports the model-scale virus RNA indicator *υ*(*t*) under the same epidemic modeling assumptions described for the hospitalization indicator *h*(*t*). Given the transmission model, the timing of both the virus RNA concentration and hospitalization indicators is clear, and provides a clue as to what might be expected when examining the timing of observed hospital admissions and SARS-CoV-2 data in sewage sludge. We turn to such an empirical analysis in the next section.

## 4 Scaling Indicators to Transmission: Hospital Admissions and SARS-CoV-2 RNA in Sewage Sludge

Consider a model-scale infection indicator *y*(*t*) as earlier described, and let *Y*(*t*) be the random variable denoting the real-world-scale observable value of this indicator at time *t*. For example, corresponding to the model-scale hospitalization indicator *h*(*t*), the real-world number of hospital admissions observed on day *t* is the random variable *H*(*t*). Similarly, random variable *V*(*t*) denotes the actual concentration of RNA observed in sewage sludge on day *t*, corresponding with the model-scale virus RNA concentration indicator *υ*(*t*).

The observable indicator *Y*(*t*) is modeled as a random variable with mean proportional to *y*(*t*), that is,

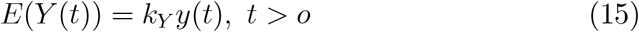

for some indicator-specific constant *k_Y_*. We thus scale observable indicators to their model-scale values in expectation. Note from equation (11) that

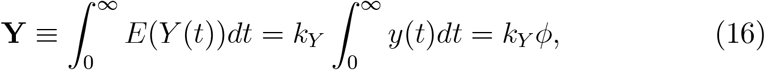

a result we will exploit in Section 6 below.

We also allow the indicator variance 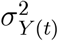 to depend upon *y*(*t*). Given the transmission model, we treat *Y*(*t*) as conditionally independent of *Y*(*t*′) for all *t ≠ t′*, for correlation in observed values across time would almost entirely be due to the underlying epidemic. The specific probability law presumed for *Y*(*t*) given *y*(*t*) can differ by infection indicator, as will become clear by example. Given observed indicator values at different points in time and an underlying epidemic model, one can estimate the scaling constants *k_Y_* (and variance parameters if needed) via maximum likelihood or other methods.

The Peccia et al (2020) study of sewage sludge obtained daily COVID-19 admissions data to the Yale New Haven Hospital restricted to residents of the same four Connecticut towns served by the local wastewater treatment plant. The data record the first such admission as occurring on March 14, 2020, 24 days following our February 19 starting date (*t* = 0). We focus here on daily admissions data recorded through May 1, 2020 (*t* = 72).

Daily hospital admissions data *h_t_* are modeled as realizations of a Poisson random variable *H*(*t*) with mean proportional to the model-scale indicator *h*(*t*) developed earlier, that is,

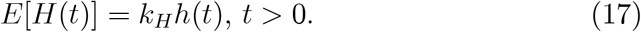

The Poisson log likelihood corresponding to the hospital admissions data covering March 14 (*t = 24*) to May 1 (*t = 72*), 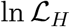, is thus given by

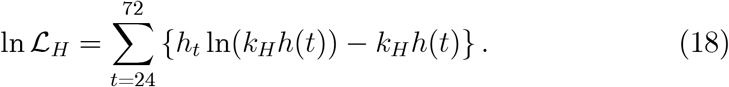

Also as reported in Peccia et al (2020), sludge samples from the local wastewater treatment plant were tested for SARS-CoV-2 RNA concentrations with two different primers applied to two sample replications daily. These values were adjusted to control for day to day variations in treatment plant flow, sludge solids content, and RNA extraction efficiency (Peccia et al 2020). The data we employ here are *υ_t_*, the day *t* average of these four adjusted values with measurement units 10^5^ SARS-CoV-2 RNA copies / ml sludge. We again focus on data collected from March 19 through May 1 (*t = 29,…, 72*) for a total of 44 daily observations. We model *υ_t_* as realizations of a Normal random variable *V*(*t*) with mean *E*[*V*(*t*)] = *k_V_υ*(*t*) and variance 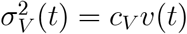 to allow for over- or under-dispersion relative to the mean^1^. The Normal log likelihood corresponding to the sludge data, 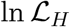, thus equals

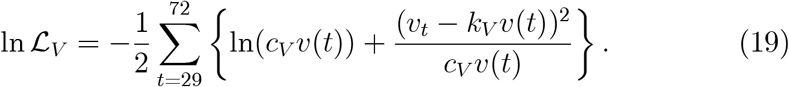

We estimate five parameters from the hospital admissions and sludge data via maximum likelihood, conditional upon the epidemic model (which implies the forward generation lag density 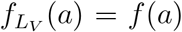 based on Li et al (2020), and hospital lag density 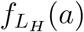 based on Lewnard et al (2020)). Three of the parameters estimated are the hospitalization scaling constant *k_H_*, the sludge RNA scaling constant *k_V_*, and the sludge RNA variance scaling constant *c_V_*. The fourth parameter estimated is *R*_0_ which scales the strength of the outbreak and enables direct comparison to SARS-CoV-2 epidemics elsewhere. The final parameter estimated is *π*(0), which sets the initial condition of the model via the relation

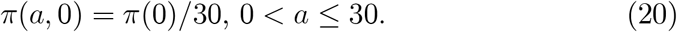

This modeling choice reflects the random arrival of imported infections to the area of study in the thirty days preceding the onset of community transmission, in effect determining the placement of the main epidemic wave without changing its shape. A larger value of *π*(0) would pull the epidemic earlier in time, while a smaller value would push the epidemic later. In this way, the hospital admissions and sludge virus RNA concentration data jointly determine the size and the placement of the epidemic wave while impacting the transmission dynamics via the model described in equations (4-7). Consequently, population susceptibility at time 0 is given by *s*(0) = 1 − *π*(0).

Table 1 reports the maximum likelihood estimates and standard errors computed by inverting the Hessian matrix of the log likelihood function (Cox and Hinkley 1974) following maximization of 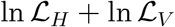, while the fit of the observed data to the scaled model indicators is illustrated in Figures 2 and 3, which plot 95% prediction intervals about the expected indicator values in addition to the data. The hospital admissions data are plotted in Figure 2. Though noisy, the admissions data correspond to the modeled pace of the epidemic, with most values falling within the 95% prediction intervals.

Figure 3 reports the observed and modeled SARS-CoV-2 RNA concentrations (in 10^5^ RNA copies / ml sludge) from the sewage study along with conservative 95% prediction intervals. While peak RNA virus concentrations are higher than what would be expected based on the model, the data again match the estimated pace of the epidemic, suggesting that community virus RNA concentration in sewage sludge can indeed be represented by the transmission potential in an epidemic model. The data rise and fall as expected, albeit with much random noise to be sure.

Note that the estimated reproductive number is 2.38 with a 95% confidence interval ranging from 2.18 to 2.58. This places the local SARS-CoV-2 outbreak in New Haven squarely in the middle of reproductive numbers estimated elsewhere (for examples see CDC (2020) and MIDAS (2020)). What is noteworthy is that this reproductive number was estimated from a model linking transmission to hospital admissions and SARS-CoV-2 RNA concentrations measured in sewage sludge. The data, not the model, determined the magnitude of *R*_0_, supporting the plausibility of the hospitalization and generation time lag distributions employed to match the observed data to an underlying transmission model.

Together with the epidemic model, these data help explain one of the findings in the Peccia et al (2020) study, which is that the SARS-CoV-2 RNA signal from the sewage sludge led hospital admissions by only 4 days, when many were expecting a much earlier signal. The model shows that the natural time lag for virus RNA concentration is governed by the mean forward generation time, estimated at 8.9 days in this model. Given an average 13.5 day lag from infection to hospitalization documented elsewhere (Lewnard et al 2020), tracking the outbreak by relying on the sewage sludge RNA signal leads similar tracking by hospital admissions by 4.6 days on average, which is very close to the purely statistical time series results reported by Peccia et al (2020).

## 5 Sensitivity Analyses

The analysis of Section 4 relies on two particular lag distributions: the forward generation time based on Li et al (2020) and the hospital admissions density based on Lewnard et al (2020). Different lag distributions could generate different results yet also appear reasonably consistent with the data. In this section we will summarize maximum likelihood scalings as in the previous section but using alternative lag density combinations.

Starting with the forward generation time density that is used to both drive the epidemic model and provide a model-scale indicator for SARS-CoV-2 RNA in sewage sludge, we turn to the meta-analysis of several published studies reported by Park et al (2020). The consensus distribution from that analysis is also a gamma density but with a mean (standard deviation) of 8.5 (6.1) days. Figure 4a plots both the Li et al (2020) and Park et al (2020) generation time densities, from which one can see that the timing of transmission is relatively early under the Park et al (2020) model relative to our base case of Li et al (2020). Regarding the distribution of the time from infection until hospital admission for those requiring hospitalization, CDC (2020) recommends a mean of 12 days based upon 6 day mean times from infection until the onset of symptoms, and from the onset of symptoms until hospitalization. We model this “short” hospitalization lag as a gamma distribution with a mean (standard deviation) of 12 (6) days. To explore the possibility that sludge RNA provides a longer lead time over COVID-19 hospitalizations, we also consider a “long” hospitalization gamma-distributed lag with a mean (standard deviation) of 15 (6.7) days. Figure 4b plots the short, base case (following Lewnard et al 2020), and long hospitalization lag densities.

Table 2 reports the mean lead time (given by the difference between the mean hospitalization and forward generation/sludge RNA lags), log likelihood function, estimated reproductive number *R*_0_ and estimated initial fraction infected *π*_0_ for all six combinations of the hospitalization and forward generation lags. While all the maximized log likelihood values are comparable, the late transmission Li et al (2020) generation time density fits slightly better than the early transmission Park et al (2020) generation time density for all three hospital lags, while the hospitalization lag densities fit best from short to Lewnard et al (2020) to long for both generation time densities. The point estimates for *R*_0_ range from 2.2 (early transmission and long hospitalization lag) to 2.43 (late transmission and short hospitalization lag), and are all within the 95% confidence interval provided by our earlier base case analysis in Section 4. The point estimates for the initial fraction infected *π*_0_ range from 0.010 to 0.019, and are also all within the 95% confidence interval estimated in our base case. Examining the log likelihood values, there is slightly more evidence favoring shorter RNA sludge lead times. The best fitting model (late transmission and short hospital lag) estimates that sludge RNA provides an expected lead time of only 3.1 days over hospital admissions data, while the worst fitting model (early transmission and long hospital lag) has an expected lead time of 6.5 days. Together these results suggest that the sludge RNA signal provides a 3 to 5 day lead time over hospital admissions, consistent with what was found based on statistical time series analysis in Peccia et al (2020).

**Table 2.**

| **Generation Time** | **Hospital Lag** | **Lead Time** | **Log Likelihood** | $R_0$ | $\pi_0$ |
| --- | --- | --- | --- | --- | --- |
| Late | Short | 3.1 | 1332.8 | 2.43 | 0.013 |
| Early | Short | 3.5 | 1332.3 | 2.26 | 0.010 |
| Late | Base Case | 4.6 | 1331.1 | 2.38 | 0.016 |
| Early | Base Case | 5.0 | 1330.2 | 2.22 | 0.013 |
| Late | Long | 6.1 | 1329.5 | 2.36 | 0.019 |
| Early | Long | 6.5 | 1328.4 | 2.20 | 0.015 |

## 6 Epidemic Insights

The epidemic model developed describes an unmitigated outbreak among a population at risk for SARS-CoV-2 infection, but in Connecticut where this study took place, social distancing and lockdown-like stay-at-home orders were imposed on March 22 (Lamont 2020a) and extended past the duration of our study period (Lamont 2020b). An unmitigated outbreak with *R*_0_ = 2.38 would leave 87.5% of the population infected, but clearly that did not occur in the greater New Haven urban area.

Nonetheless, the timing of the sludge and hospital admissions data are consistent with an unmitigated outbreak. Models reflecting the effect of lockdowns and social distancing applied uniformly to large populations show that transmission is both delayed and slowed, resulting in the oft-cited flattening of the epidemic curve (Ferguson et al 2020, Kissler et al 2020, Kaplan 2020b). The epidemic models we have shown to be consistent with the observed data do not behave this way, suggesting that the effect of the stay-at-home orders was not experienced uniformly. A different possibility is that stay-at-home restrictions essentially bifurcated the population into two groups: a large group of citizens whose compliance with stay-at-home restrictions removed them from potentially infectious interactions, and a smaller group of essential workers, other vulnerable persons such as nursing home residents, or non-compliant individuals that, due to necessity or choice, continued to experience exposures via interactions with others, enabling continued transmission. Were that the case, then members of the “exposed” population could have experienced an unmitigated outbreak while compliant individuals escaped unscathed. The epidemic model of this paper might only portray transmission among the exposed population, yet all infections and hence hospitalizations and SARS-CoV-2 RNA in the sewage sludge would have emanated from this group.

To investigate this possibility, we will use our model results to produce a back-of-the-envelope estimate of the fraction of the population that was exposed, and consequentially the fraction of the entire population that complied with the stay-at-home restrictions. We will also estimate the implied number of infections that must have occurred under this bifurcation hypothesis, including the initial number of infected persons circa February 19, 2020 (which recall is clock time 0).

First, we apply equation (16) to the expected total number of hospital admissions over all time from this outbreak which yields

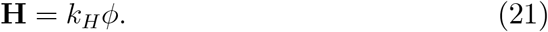

Next, define **N** and **C** as the number of persons in the exposed population and the total number of diagnosed COVID-19 cases that occurred in that population respectively over all time. A second equation for **H** is given by

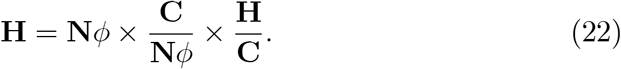

Equating equations (21) and (22) yields

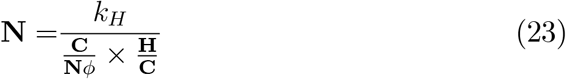

as our estimate for the size of the exposed population. Focusing on the denominator, the ratio **C**/**N***ϕ* is the ratio of diagnosed COVID-19 cases to infections in the exposed population, also known as the case ascertainment ratio. Under the bifurcation hypothesis, this will be the same as the ratio of diagnosed COVID-19 cases to infections in the overall population, as infections and cases only accrue in the exposed group. A direct estimate of the reciprocal of the case ascertainment ratio (that is, the number of infections per diagnosed COVID-19 case) was reported by Havers et al (2020) for Connecticut covering March 23-May 12 of 2020 viaapopulation-based seroprevalence survey. Havers et al (2020) estimated an average of 6.0 infections per case (95% confidence interval 4.3–7.8). The ratio **H**/**C** can be estimated directly from Peccia et al (2020) who reported that there were 2,674 diagnosed cases and 734 hospital admissions emanating from the four towns served by the local wastewater treatment plant. The constant *k_H_* was estimated in our base case analysis of Section 4 to equal 1006.6 (see Table 1).

Substituting the values above into equation (23) we obtain

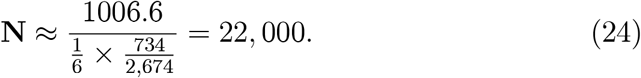

Recalling that 200,000 persons are served by the local wastewater treatment plant, we estimate that 22, 000/200, 000 = 11% of the population were exposed to infection while the stay-at-home restrictions protected the remaining 89% of the population from infection, indicating a high level of compliance with the public health regulations. Further insight can be gained using equation (8) and our base case estimate of *R*_0_ = 2.38 to estimate that the fraction of the exposed population infected over all time in this outbreak is given by *ϕ* = 0.875. However, as of May 1 (day 72) when this study concluded, the epidemic model suggests that only 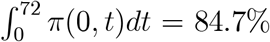 of the exposed population had been infected, which means that 22, 000 × 0.847 ≈ 18, 600 persons were infected by May 1 in the New Haven metropolitan area (or 9.3% of the total population). As an independent check, Havers et al’s (2020) serological estimate for the ratio of infections to cases suggests that given 2,674 diagnosed cases, a 95% confidence interval estimate for the number infected runs from 11,500 to 20,900 persons, in agreement with our model-based result. Finally, given our estimate that 1.6% of the exposed population was already infected as of February 19, 2020 (*π*_0_; see Table 1), we estimate that *22*,000 × 0.016 ≈ 350 persons were already infected at that early date.

## 7 Summary

This paper has focused on modeling lagging epidemic indicators and how they relate to each other. The approach has been to utilize an epidemic model as a basis for scaling indicators like hospital admissions or SARS-CoV-2 RNA observed in sewage sludge. After characterizing how indicators lag incidence, we showed how one could use an epidemic model to simultaneously estimate the placement of an epidemic wave (via estimating the initial condition), the strength of an outbreak (via estimating *R*_0_), and situate lagging indicators appropriately, allowing one to view the data in a more epidemiologically meaningful way. Using data from a recently published study of SARS-CoV-2 RNA concentrations observed in municipal sewage sludge, we showed why the RNA data were only able to shorten the time from infection to signal by 3 to 5 days relative to hospital admissions. The RNA and hospitalization data jointly implied an epidemic with *R*_0_ of approximately 2.38, well within the range implied by numerous studies. To reconcile this finding with the fact that Connecticut was under strict lockdown-like stay-at-home orders throughout most of the study period, we postulated that the stay-at-home restrictions effectively bifurcated the population, resulting in an unmitigated outbreak among an estimated 11% of the population who remained exposed to infections while sparing the remaining 89% who complied with the restrictions. Overall we estimated that about 9.3% of the total population became infected. To our knowledge, ours is the first study to develop such population-level findings based on exploiting the infection signal contained in SARS-CoV-2 RNA in sewage sludge and COVID-19 hospital admissions data.

## Data Availability

Data reported in
https://www.medrxiv.org/content/10.1101/2020.05.19.20105999v2

## Acknowledgements

The authors thank Doug Brackney, Arnau Casanovas-Massana, Nate Grubaugh, Albert Ko, Saad Omer and Dan Weinberger for comments on an earlier draft of this manuscript.

## Footnotes

1 We also considered a model with constant variance but the model did not fit the data as well.

